# Challenges Facing Early-Career Physician–Scientists in the United States Amid Recent Policy Shifts: Findings from a National Survey

**DOI:** 10.64898/2026.04.26.26351791

**Authors:** Abdelrahman Abushouk, Aleksander Obradovic, Alisha Faraz, Aisha Siebert, Han Naung Tun, Evan Noch, Jennifer M Kwan

## Abstract

**Background:** Amid persistent structural barriers and recent national policy changes, early-career physician–scientists face mounting challenges that threaten the sustainability of the biomedical research pipeline in the United States.

**Methods:** We surveyed early career physician-scientists collecting demographic data, career development support, distribution of clinical and research responsibilities, funding, and perceived career challenges. The survey was distributed by email to the department chairs at 110 institutions in the United States.

**Results:** A total of 175 surveys were completed. About half 50.8% (n=89) of respondents received a career development award, with 28.9% of respondents reporting limited institutional/departmental support. The most reported challenges were balancing clinical, research, and educational responsibilities (72.5%, n=127); balancing work and family responsibilities (48%, n= 84); limited funding opportunities (48%, n=84); and under-compensation (34.3%, n=60). About 57.7% (n=101) of respondents had considered leaving academic medicine within the next two years, and 83.2% (n=139) indicated a >50% likelihood of doing so within five years. The most frequently cited reasons for attrition were funding challenges (72%, n=126), under-compensation (42.3%, n=74), feeling unhappy or stressed (40.6%, n=71), and burnout (37.7%, n=66). Furthermore, 43.9% (n=76) of respondents reported considering relocation outside the United States for better academic working conditions, and 10.4% (n=18) had already been contacted by institutions abroad.

**Conclusion:** Early-career physician–scientists face substantial structural and financial challenges, with limited institutional support, high rates of burnout, and widespread intent to leave academia. These findings underscore an urgent need for sustained investment, targeted retention strategies, and policy reforms to stabilize and strengthen the physician–scientist workforce in the United States.

## Introduction

Physician–scientists have long been a driving force in translating scientific discoveries into meaningful advances in patient care.^1^ They are healthcare professionals who dedicate a substantial portion of their career to research while maintaining active engagement in clinical practice.^2^ This community encompasses a wide spectrum of investigators, from clinicians with strong research skills who conduct independent investigations to those with formal dual-degree training (MD–PhD) designed to integrate clinical expertise with rigorous scientific inquiry.^3^ Serving as a critical bridge between the laboratory and the bedside, this workforce identifies clinically relevant questions and translates them into discoveries that improve patient outcomes across disciplines.^4^

However, the physician-scientist pipeline has long been laden with challenges, affecting the community at every training stage, particularly those in the early phases of training.^5^ Consequently, this scientific workforce has markedly contracted, declining from approximately 4.5% of the medical community in the 1980s to about 1.5% over the last decade.^6^ In addition, numerous studies have reported data consistent with the ongoing workforce contraction, characterized by a markedly aging demographic and a decline in the number of early-career investigators.^7,8^ This trend is further compounded by decreasing interest among medical students in pursuing research careers. Additionally, our recent survey revealed that 49% of early-career physician–scientists have contemplated leaving their academic positions.^1,9^ This decline reflects not only persistent structural barriers but also insufficient institutional and policy support that aim to improve retentions of these skilled individuals.^5^ In this context, policy changes implemented across the nation last year led to the termination of 694 NIH grants across 24 of the 26 institutes and centers, with a cumulative loss of $1.81 billion in research funding between February and April 2025.^10^ According to new data from the NIH, the funding rate for early-career investigators dropped from 26% in 2024 to 19% in 2025.^11,12^

Historically, stakeholders have shed light on the importance of sustained investment through institutional commitment and grant mechanisms, which remain a cornerstone for preserving the vitality of the workforce amid growing professional pressures and evolving policy landscapes that threaten to impede scientific progress.^1^ These concerted efforts have led to the establishment of organizations that provide platforms for advocacy and support for physician-scientists, including the American Physician Scientists Association (APSA) for MD PhD students and the American Junior Investigator Association (AJIA) for early career investigators.^13,14^ In this paper, we provide an overview of key challenges facing early-career physician-scientists in the United States, along with their perspective amid recent policy changes and broader issues affecting the biomedical and clinical research community.

## Methods

The survey instrument collected demographic information (age, training level, gender, race, medical specialty, and geographic region) and responses related to career development support, distribution of clinical and research responsibilities, funding, and perceived career challenges. These challenges included the likelihood of leaving academic medicine and the reasons for such decisions. The survey was distributed by email to department chairs at 110 U.S. institutions, who were asked to share it with their early career investigators. The complete survey tool is available in the supplementary materials (**Supp Table 1**).

The final analytic sample included 175 participants, after exclusion of incomplete responses with missing demographic data. Statistical analyses were conducted by first assessing pairwise associations between each demographic variable and binarized survey response variable using Fisher’s exact test. To account for multiple comparisons, p-values were adjusted using the Benjamini–Hochberg false discovery rate (FDR) correction. Statistical analyses were performed in R v4.2.3 programming language in RStudio v2024.04.1 integrated development environment.

## Results

A total of 175 complete survey responses were included in the analysis. Among respondents, 78 (45.3%) identified as female, and the largest age group was 35–44 years (n= 106; 62.7%). Most respondents identified as White (n=112; 64.7%), followed by East Asian (China, Japan, and Korean; n=21; 12.1%) and African American (n=13; 7.5%), while only 10 (5.8%) identified as Hispanic. The respondents represented a broad range of academic ranks. Assistant professors comprised the largest group (n=82; 46.6%), followed by residents (n=25; 14.3%) and fellows (n=21; 12%). Participants spanned multiple medical domains, predominantly general medicine and related subspecialties (n=101; 57.7%), neurology/psychiatry (n=18; 10.3%) pathology (n=11; 6.3%), and surgical subspecialties (n=8; 4.6%). Geographic distribution showed the greatest representation from the Northeast (n=61; 35.7%), followed by the Southwest (n=39; 22.8%), Midwest (n=29; 17%), Southeast (n=24; 14%), and Northwest (n=11; 6.4%). The demographic characteristics are illustrated in **Table 1** and heatmap for geographic representation of respondents is represented in **Figure 1**.

**Figure 1:**
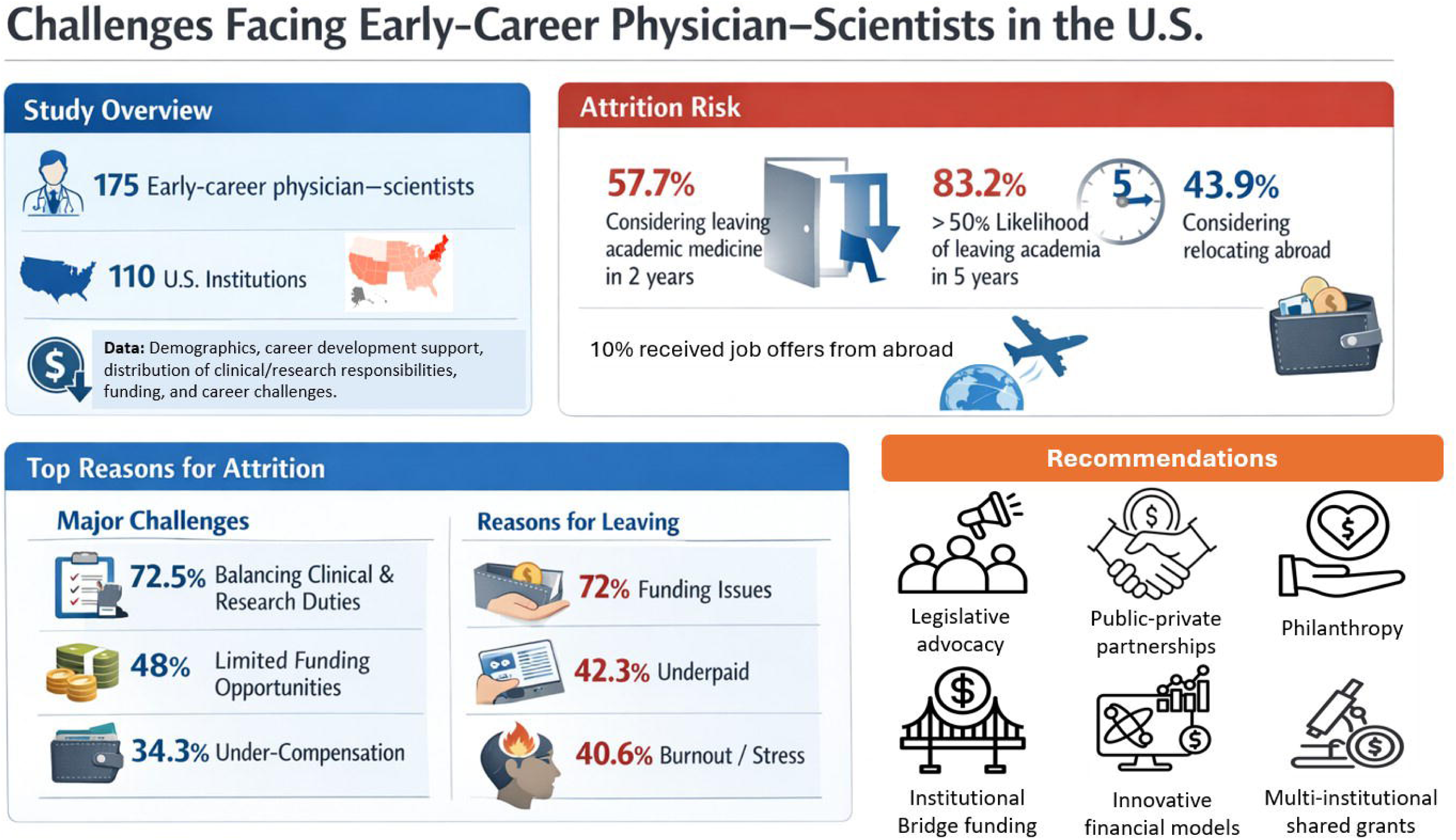
Heatmap representation of geographic distribution of survey respondents.

**Table 1:** Demographics of the study participants.

| Characteristic | N= 175 (100%) |
| --- | --- |
|  | n (%) |
| Age years (N=169) |  |
| 25-34 | 35 (20.7) |
| 35-44 | 106 (62.7) |
| 45-54 | 18 (10.7) |
| 55-64 | 4 (2.4) |
| 65+ | 6 (3.6) |
| Male Gender (N=172) |  |
| Male | 90 (52.3) |
| Female | 78 (45.3) |
| Other | 2 (1.2) |
| Prefer not to describe | 2 (1.2) |
| Race (N= 173) |  |
| White | 112 (64.7) |
| African American | 13 (7.5) |
| East Asia | 21 (12.1) |
| Other | 27 (15.7) |
| Hispanic (N= 173) | 10 (5.8) |
| Academic rank (N=175) |  |
| Resident | 25 (14.3) |
| Fellow | 21 (12) |
| Instructor | 16 (9.1) |
| Assistant Professor | 82 (46.6) |
| Associate/Full Professor | 26 (14.9) |
| Other | 5 (2.9) |
| Clinical Specialty |  |
| Internal Medicine/subspecialties | 101 (57.7) |
| Neurology/Psychiatry | 18 (10.3) |
| Surgical Specialties | 8 (4.6) |
| Pathology | 11 (6.3) |
| Other | 37 (21.1) |
| Geographic distribution (N=171) |  |
| Northeast | 61 (35.7) |
| Northwest | 11 (6.4) |
| Midwest | 29 (17) |
| South | 6 (3.5) |
| Southeast | 24 (14) |
| Southwest | 39 (22.8) |
| Puerto Rico | 1 (0.6) |

Data represents n (%)

Furthermore, respondents reported varied research-to-clinical effort ratios. About 74.2% (121/163) of the sampled participants reported that they dedicated 50% or more of their time to research, with 7 (4.3%) reporting full time research. Notably, among those with a research effort of less than 50%, 81.8% wished to dedicate more time to research. When asked about what area of research the participants intended to spend most of their professional time, 45 (25.7%) chose basic, 53 (30.3%) translational, and 44 (25.1%) clinical research.

In terms of grant funding, more than half of respondents (89/175; 50.8%) reported having received a career development award (CDA), including K08 (n=36), K23 (n=11), and R01 (n=16). **Figure 2** shows the proportions of respondents who did not have a CDA by different career stages. There was no significant difference in the proportion of those who received a CDA vs not amongst respondents who indicated wanting to the leave their research careers (58.9% vs 50.6%, p=0.374) or the country (45.6% vs 38.8, p=0.434). Among the 80 respondents who provided information on submission attempts, 36 (45%) secured funding on their first attempt, 31 (38.8%) on their second, while the remaining participants took three or more attempts. In addition, institutional support was limited. Only 50/171 (29.2%) reported base salary equalization, and 50/173 (28.9%) reported research-related incentives or RVU adjustments. Among those 50 participants, 28 (56%) reported protected time tied to grant activity, 25 (50%) reported bonus compensation for funded grants, 16 (32%) reported reduced clinical or administrative responsibilities, while only one reported RVU credit for grant submissions (**Table 2)**.

**Figure 2:**
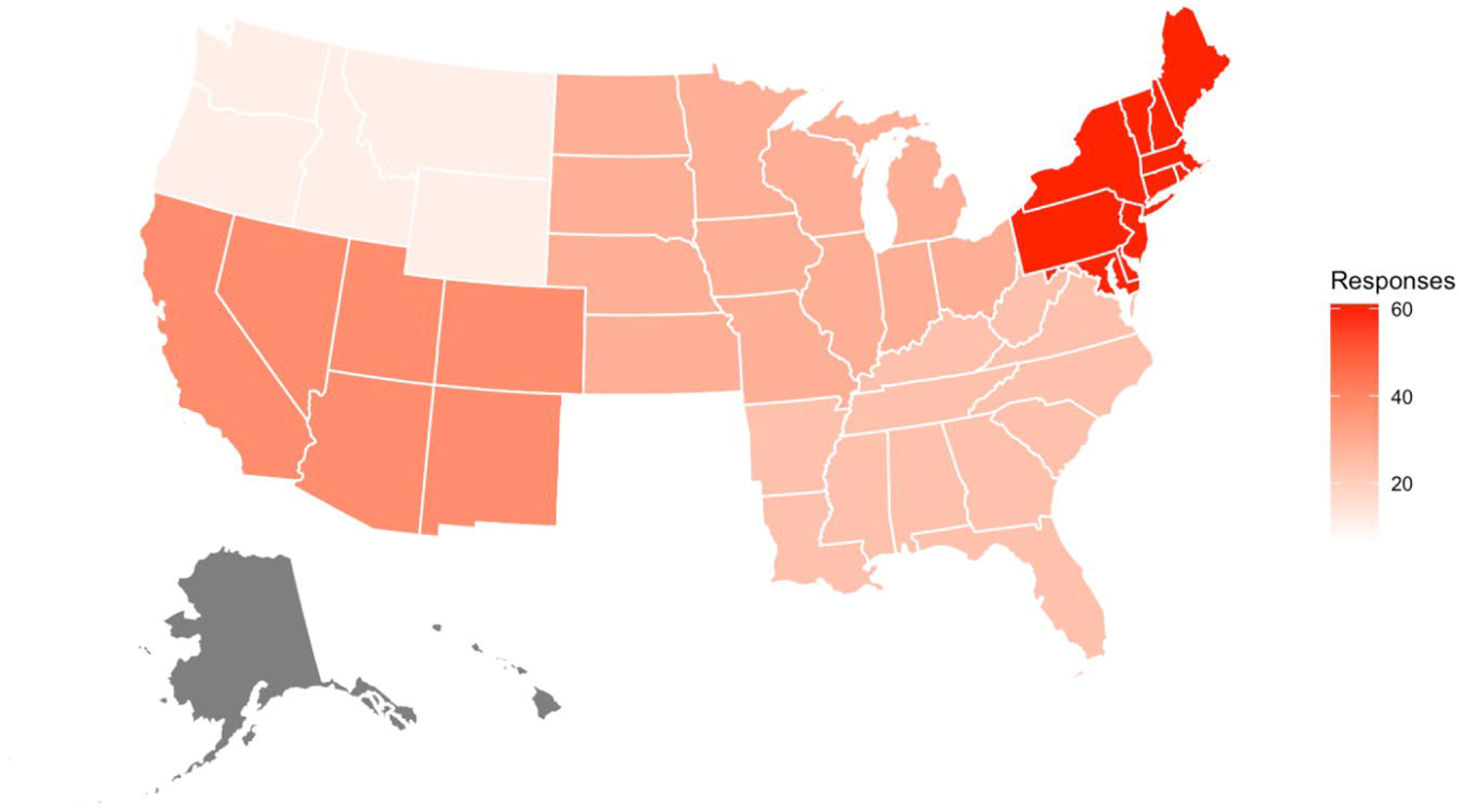
Comparison of those who have CDA vs no CDA on intention of leaving research or the country and stage in training with CDA.

**Table 2:** Current research landscape and funding mechanisms among the participant early physician-scientist population.

|  | n (%) |
| --- | --- |
| Current Research Time Commitment (N=163) |  |
| ≥ 50% | 121 (74.2) |
| ≥ 75% | 92 (56.4) |
| 100% | 7 (4.3) |
| Research Area |  |
| Basic | 45 (25.7) |
| Translational | 53 (30.3) |
| Clinical | 44 (25.1) |
| Other | 33 (18.9) |
| Having a Career-development award (N=175) | 89 (50.8) |
| K08 | 36 (20.6) |
| K23 | 11 (6.3) |
| R01 | 16 (9.1) |
| Other K awards (K12, KL2, etc.) | 35 (20) |
| Departmental/Institutional Support |  |
| Base salary equalization (N= 171) | 50 (29.2) |
| Other research incentives (N= 173) | 50 (28.9) |
| Protected time tied to grant activity | 28 (16) |
| Bonus compensation for funded grants | 25 (14.3) |
| Reduced clinical/ administrative responsibilities | 16 (9.1) |
| RVU Credit for grant submissions | 1 (0.6) |

Data represents n (%)
If N is not included, the total number of respondents = 175

The most reported challenges by our respondents when asking about their early career/faculty transitions were balancing clinical, research, and educational responsibilities (n=127; 72.5%); balancing work and family responsibilities (n=84; 48%); limited funding opportunities (n= 84; 48%); and under-compensation (n=60; 34.3%). More than half of respondents (n=101; 57.7%) had considered leaving academic medicine within the next two years, and 139 (83.2%) indicated a >50% likelihood of doing so within five years. No significant regional variation was observed in terms of the intent to leave academic medicine (p=0.118). The most frequently cited reasons for attrition were funding challenges (n=126; 72%), under-compensation (n=74; 42.3%), and feeling unhappy or stressed (n=71; 40.6%). Each of these factors was positively associated with the intent to leave academia: funding challenges (OR= 5.02; 95% CI: 2.46 to 10.22; p< 0.001); under-compensation (OR= 2.51; 95% CI: 1.33 TO 4.72; p= 0.004); and feeling unhappy or stressed (OR= 2.23; 95% CI: 1.18 to 4.19; p= 0.012). Furthermore, respondents expressed substantial concern about the stability of research support. The greatest concern was for the future of health disparities research (n=133; 76%), followed by vaccine-related research (n=123; 70.3%) and diversity, equity, and inclusion (DEI) research (n=110; 62.9%). Additional concerns are summarized in **Table 3**.

**Table 3:**
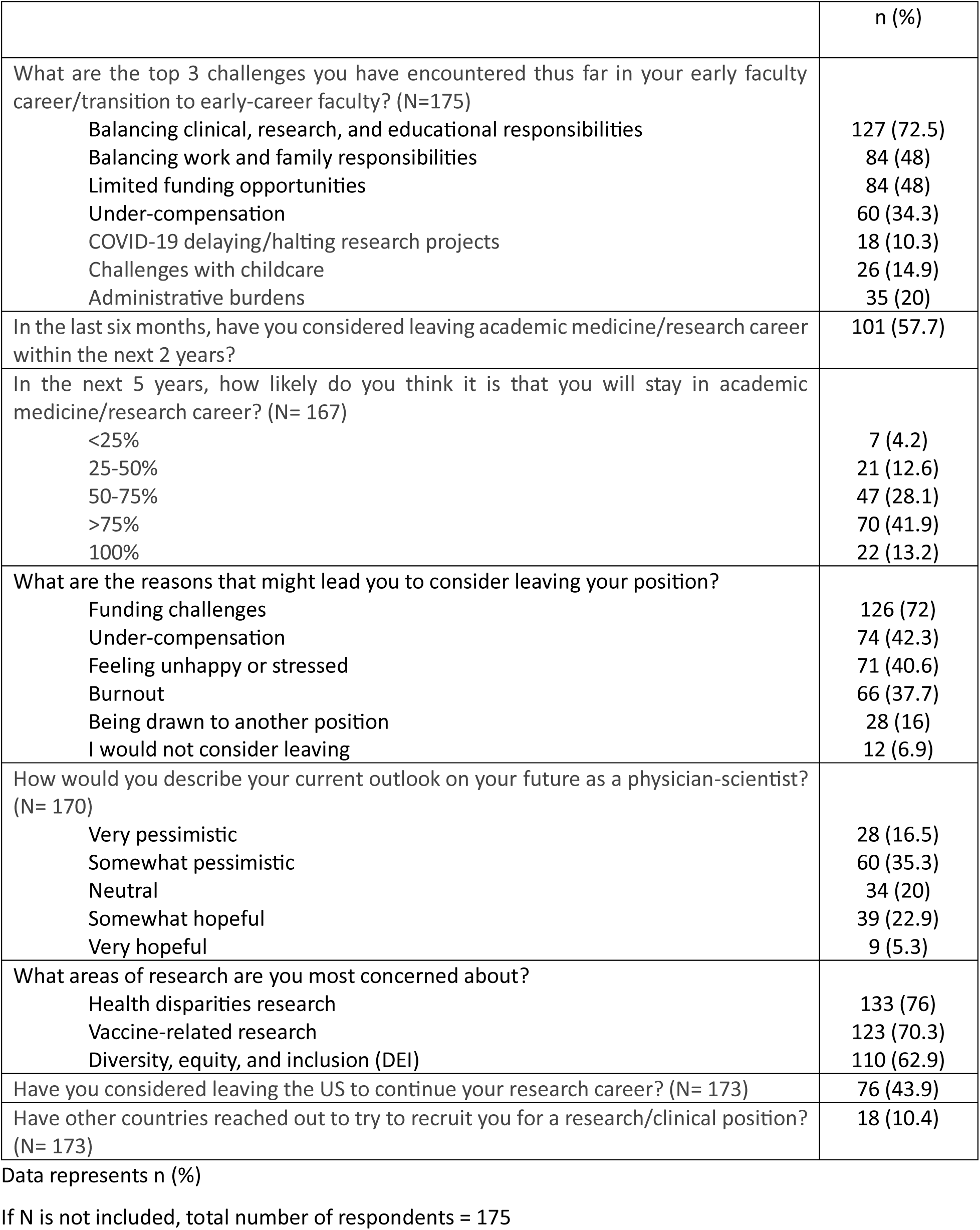
Challenges facing early-career physician-scientists and future concerns.

Taken together, these pressures influenced their outlook on academic medicine: 28 (16.5%) felt very pessimistic, 60 (35.3%) somewhat pessimistic, 39 (22.9%) somewhat hopeful, and only 9 (5.3%) were very hopeful. In addition, about half of our respondents (n=76; 43.9%) reported considering relocation outside the United States for better academic working conditions and 18 (10.4%) had already been contacted by institutions abroad. We observed a regional variation in terms of the intent to leave the US, where higher percentages from the Northeast, Northwest, and Midwest expressed an intent to leave the US (p=0.027). Those considering relocation were more likely to report funding challenges (OR= 2.13, 95% CI 1.06 to 4.29; p= 0.049). Of note, no significant difference was found in terms of either leaving academic medicine (OR= 1.37; 95% CI 0.57 to 2.49; p= 0.385) or relocating outside the US (OR= 1.28; 95% CI 0.70 to 2.33; p= 0.514) between respondents who had a CDA versus those who did not. Finally, respondents identified several institutional and system-level strategies to strengthen retention and research engagement. These proposed interventions are summarized in **Figure 3**. Additional correlations between our study parameters are illustrated in **Supp Figure 1.**

**Figure 3:**
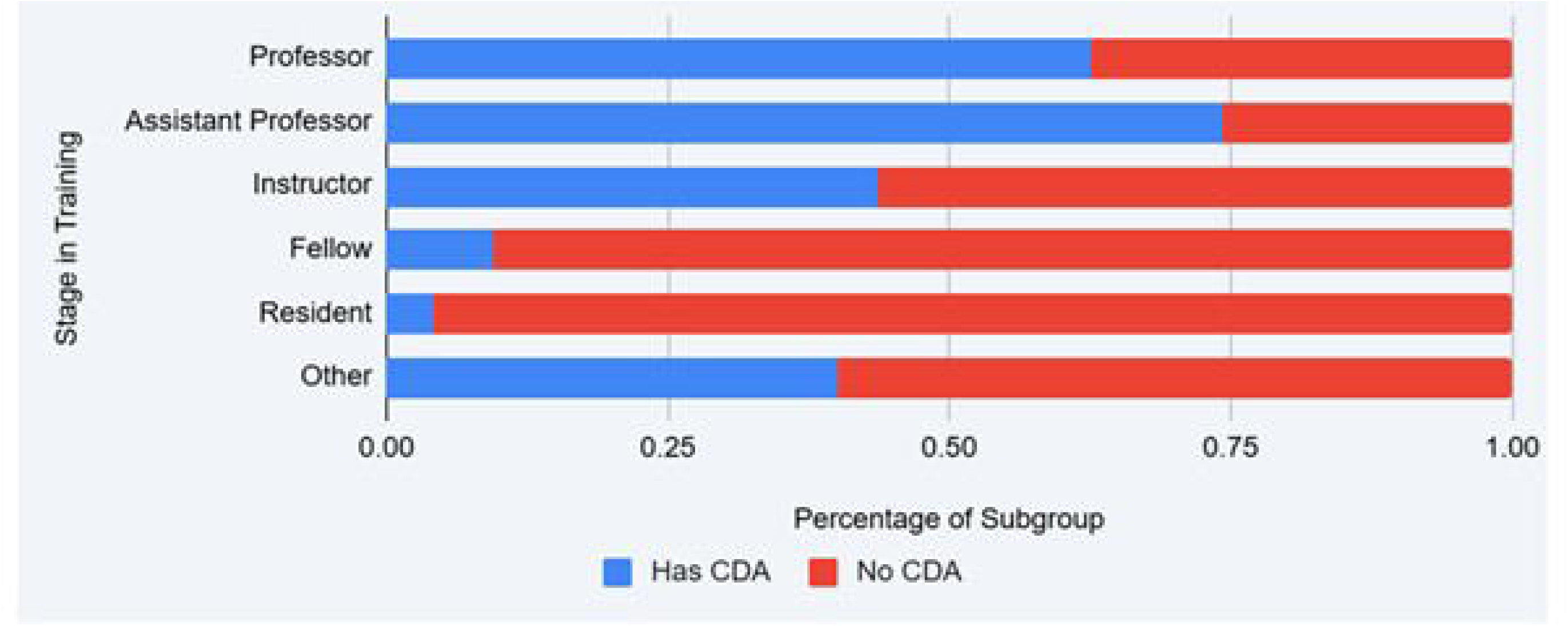
Suggestions by our study respondents to support the early physician-scientist population.

## Discussion

Our survey reveals that the physician-scientist workforce remains at continued risk of contraction. Notably, 57.7% of respondents reported having considered leaving their academic positions within the next two years. Such potential departures represent not only a loss of substantial institutional and societal investment made over many years of training, but also the forfeiture of invaluable scientific talent. When early-career physician–scientists leave academia, the burden on those who remain intensifies, perpetuating a cycle of overwork and burnout that can precipitate additional departures. This level of career uncertainty is striking, particularly as it exceeds the rate reported in a recent study from our group, in which 49% of early-career physician-scientists had considered leaving academic medicine (**Figure 4**).^9^

**Figure 4:**
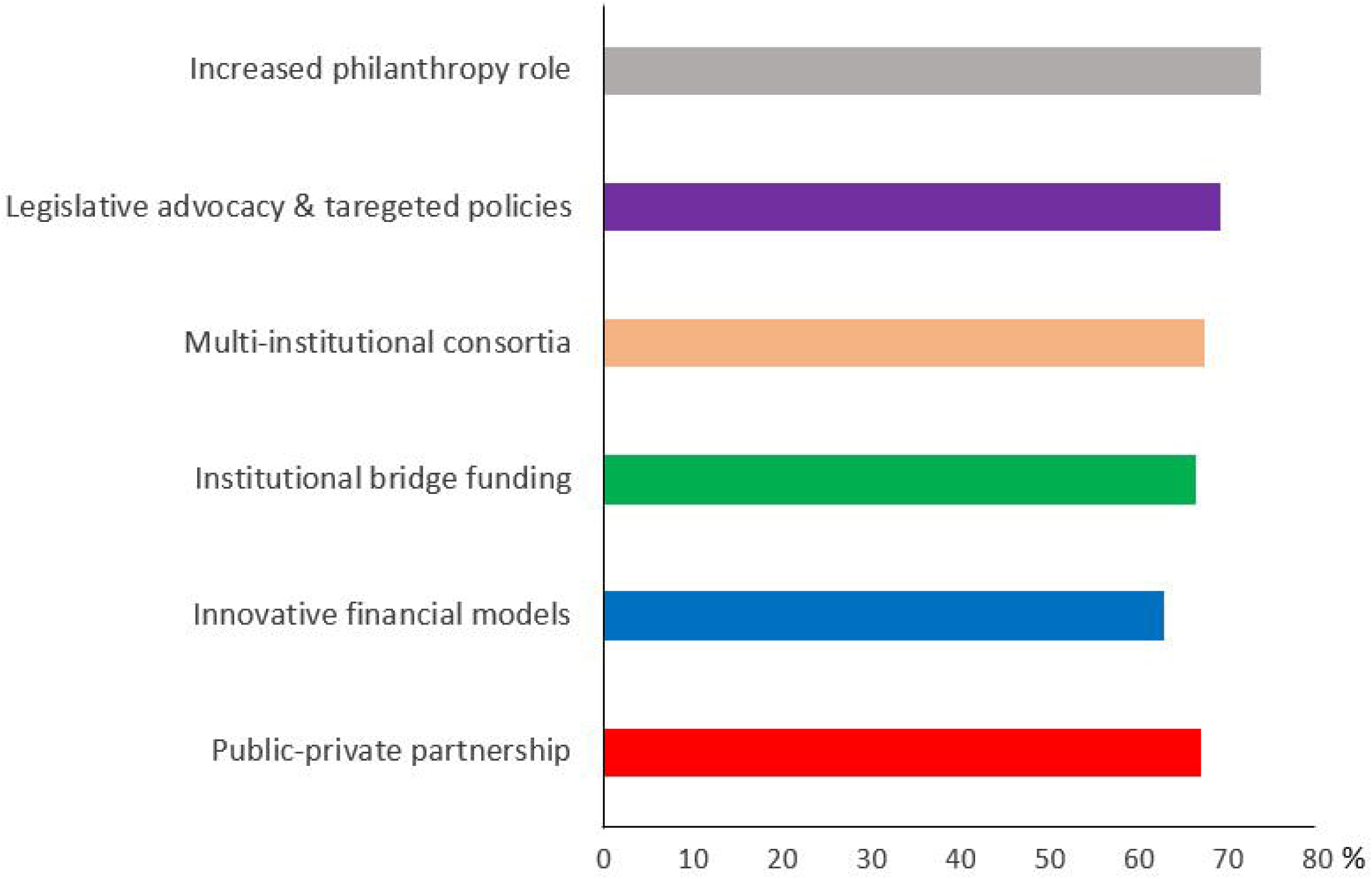
Comparison of our current survey findings to earlier survey results from our group 2024.

A sustainable funding mechanism is essential to preserve the workforce and prevent the attrition of scientists at early stages of their careers. Securing independent grant funding has always been a hurdle for early-career investigators, but our data suggest that this challenge has intensified. While a majority (51%) of respondents had obtained a CDA, among those who did, nearly half required multiple attempts to succeed. These findings align with national trends marked by increasingly competitive grant paylines and research budgets that have not kept pace with rising operational costs.^15^ These patterns underscore not only the intense competitiveness of existing funding mechanisms but also the uneven accessibility of structured research support, as evidenced by heavily skewed grant distributions across institutions, states, and stages of the pipeline. For example, 11% of NIH research funding is awarded to the top 1% of investigators, while half of all grants go to just 2% of funded organizations and 10% of states.^16^ Furthermore, a study by Lauer et al. showed that most early-career physician–scientists (72.7%) were supported by only a single research grant, placing them at high risk for funding gaps once their award cycles conclude, especially given that the success rate for securing a new research project is only 20%.^17,18^ Further, prior studies showed that funding gaps are the norm and not the exception, further exacerbating physician scientist attrition even prior to current funding challenges.^15,18^ The limited reach of these funding programs may leave many early-career investigators without the protected resources necessary to establish research independence.

Financial limitations extend far beyond funding scarcity. Many institutions lack mechanisms that help investigators balance their research with clinical obligations, such as protected research time, salary equalization, research-related incentives, or bonus compensation. As reflected in our respondents’ reports, these forms of support were notably minimal (**Table 2**). The inconsistency of such infrastructure leaves many physician–scientists effectively subsidizing their research through increased clinical effort or personal time, causing feelings of under-compensation and frustration.^9,15^ In fact, 36% of respondents identified being underpaid as a major career challenge.

Recent policy shocks have only heightened anxieties about preserving this community. In early 2025, changes in federal research priorities led to the unprecedented termination of 694 NIH grants (across 24 institutes), cutting off $1.81 billion in pledged funding within a matter of weeks.^10^ The abrupt loss of support sent shockwaves through the academic community, generating widespread uncertainty about the future of U.S. biomedical research. This impact is particularly concerning for early-career investigators, as 20% of the terminated NIH grants were early-career awards, a cornerstone source of funding that many physician–scientists rely on to launch and sustain their research careers.^10,19,20^ Our survey was conducted in the aftermath of these events, which is reflected in respondents’ expressed concerns: health disparities research was rated as the most threatened in the current climate by the majority of respondents, followed closely by vaccine-related research and studies aimed at improving diversity. Such fears are well-founded; cuts to the National Institute on Minority Health and Health Disparities and the National Institute of Allergy and Infectious Diseases, a major funder of vaccine research, accounted for the highest amount of terminated funding at nearly $506 million.^20^ In the face of this turbulent funding environment, it is understandable that even the most academically committed physician–scientists might question the viability of a long-term research career in the United States. The massive restructuring of the Department of Health and Human Services (HHS) was not limited to grant funding cuts but also included widespread layoffs at major health organizations such as the Centers for Disease Control and Prevention (CDC) and the Food and Drug Administration (FDA), which could affect their capacity to advance and regulate science, and public health, as well as undermine the trust of talented health professionals in their work environments.

These changes are not occurring in isolation. Although the United States has historically been a leader in biomedical research, recent policy shifts have contributed to a growing risk of brain drain, with many investigators considering opportunities outside the country. In our survey, 43.9% of respondents reported considering relocation for better academic working conditions, and 10.4% had already been contacted by institutions abroad, indicating active recruitment of U.S. researchers. These findings are consistent with a recent *Nature* poll in which approximately 75% of 1,650 U.S. scientists reported contemplating a move overseas in response to current federal policies, which they viewed as detrimental to early-career researchers seeking to establish independent careers.^21^ Europe and Canada were the most frequently cited destinations, reflecting perceptions of greater stability and a more supportive environment for scientific work. Importantly, our correlation analyses suggest that structural funding instability, rather than demographic characteristics, is the principal factor driving early-career physician–scientists to consider leaving academia or relocating abroad.

### Limitations

This study has some limitations. First, its survey-based design introduces the potential for selection bias. The respondent sample may not be fully representative of the broader physician–scientist community. The cross-sectional nature of the study also limits the ability to assess temporal trends or evaluate how respondents’ experiences and career intentions may evolve following ongoing funding cuts. Second, the relatively small number of respondents (n = 175) may constrain the generalizability of the findings. As observed in our previous survey analysis examining the state of early-career physician–scientists, the regional distribution of participants suggests possible overrepresentation of the Northeast and underrepresentation of the Northwest. These patterns likely reflect institutional dissemination channels and voluntary participation rather than true national proportions. Lastly, the survey instrument did not undergo formal validation. Although developed with input from relevant literature and expert review, it was not pre-tested, and no reliability assessment was conducted. This limitation may affect reproducibility and limit broader application of the instrument.

### Conclusion

Early-career physician–scientists face substantial structural and financial challenges, with limited institutional support, high rates of burnout, and widespread intent to leave academia. These findings underscore an urgent need for sustained investment, targeted retention strategies, and policy reforms to stabilize and strengthen the physician–scientist workforce in the United States.

## Supporting information

Additional correlations between our study parameters are illustrated in Supp Figure 1.

The complete survey tool is available in the supplementary materials (Supp Table 1).

## Data Availability

All data produced in the present study are available upon reasonable request to the authors

## Acknowledgements

AJIA and survey participants

## Supplementary Figure

**Supplementary Figure 1:**
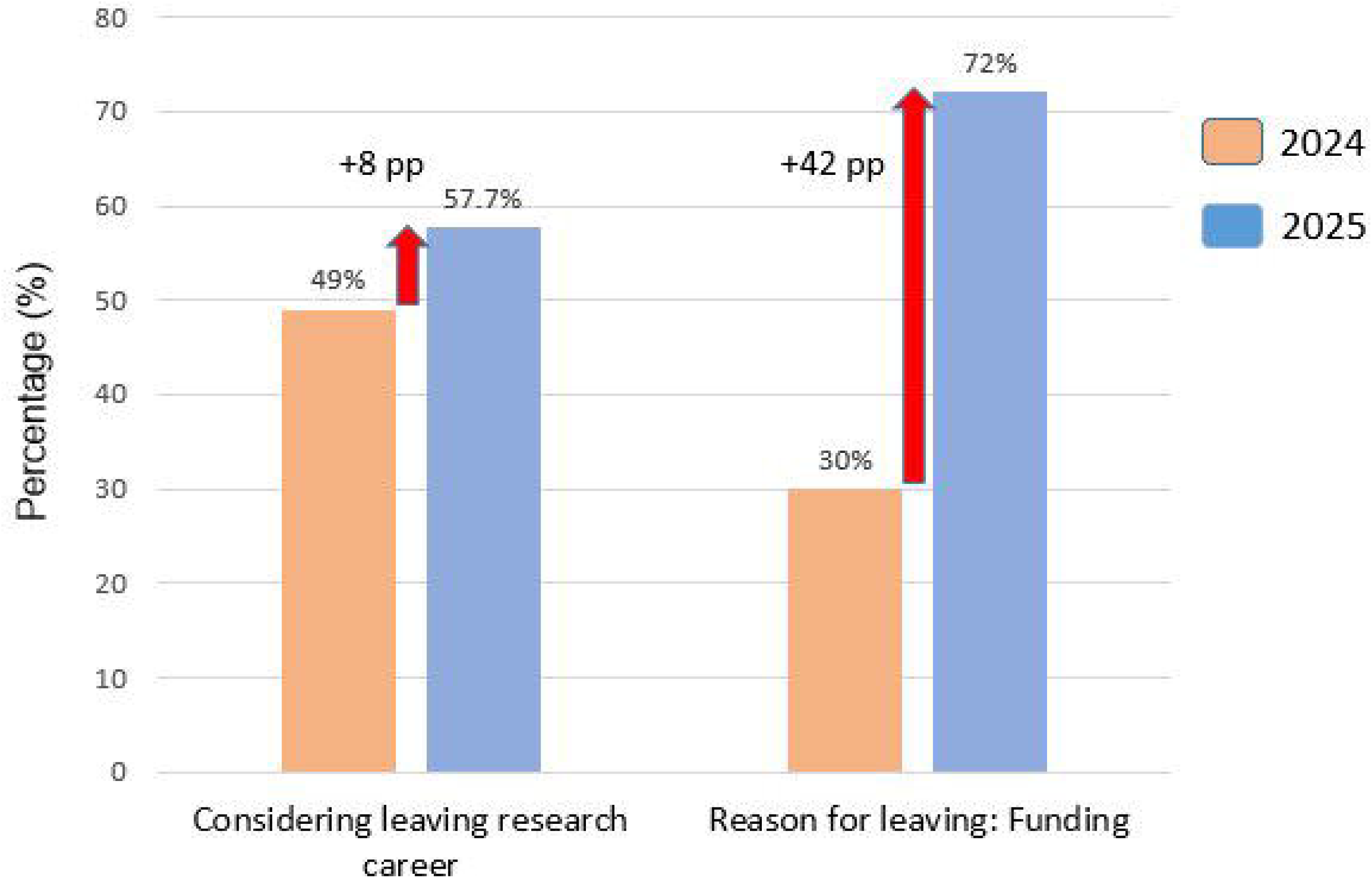
Global correlation plot between different study variables. The red boxes are filtered to statistically significant pairwise correlations after Benjamini-Hochberg multiple-testing correction

