## Supplementary figures and images for "Challenges Facing Early-Career Physician–Scientists in the United States Amid Recent Policy Shifts: Findings from a National Survey"

### Additional correlations between our study parameters are illustrated in Supp Figure 1.

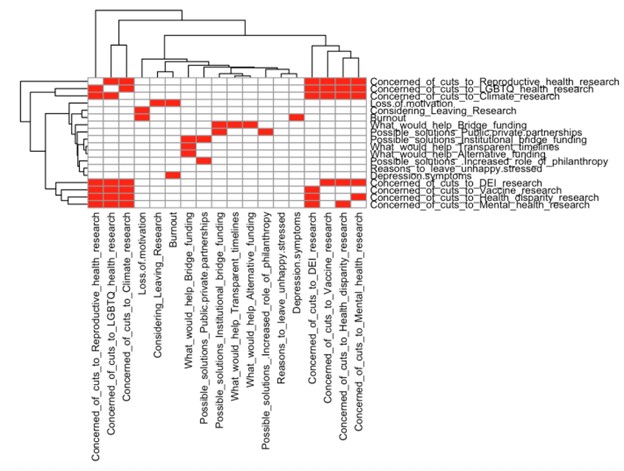
