## Supplementary material for "Challenges Facing Early-Career Physician–Scientists in the United States Amid Recent Policy Shifts: Findings from a National Survey": The complete survey tool is available in the supplementary materials (Supp Table 1).

Outlook and Support of Early Career Physician-Scientists 2025

### Consent

* 1. Physician-scientists represent only 1.5% of the biomedical workforce (Jain et al 2019) and face unique challenges. Transitioning from the resident and fellow to junior faculty career stages are the leakiest parts of the physician-scientist pipeline.

This is a survey on behalf of the American Junior Investigators Association (AJIA), a nonprofit dedicated to the success of early career physician-scientists across the full spectrum of specialties. This survey is intended to be distributed to late stage PSTP/research track residency/fellowship trainees/graduates of MD PhD and DO PhD programs and those who have graduated from PSTPs/research track residencies/fellowships within the past 10 years in the United States.

The goal of the survey is to assess how our recent research track trainees are faring, what drives their decisions in choosing faculty positions and whether recent administrative policies have impacted them.

Through the survey, we seek to characterize how early-career physician scientists have fared, important factors in choosing a career and any challenges/problems they are facing in their transition to a full time position post training. The results of the survey will be used to advocate for changes to policies/institutional structures that will help retain and advance a diverse academic medical faculty. Any significant results will potentially be published in a peer-reviewed journal.

This survey is completely anonymous and voluntary. No personally identifying information will be linked to your survey responses. There is no harm or risk associated with taking the survey.

To protect the survey taker, the account in which responses are stored is password-protected and can only be accessed by the researchers.

The survey will take approximately 7-10 minutes to complete.

If you have any questions, please feel free to contact the following research coordinators of this study: Yale School of Medicine research coordinator of this study: Jennifer M Kwan, MD PhD, the University of Texas, Southwestern Medical Center research coordinator, Evan Noch MD PhD, or the Massachusetts General Hospital research coordinator, Dania Daye MD PhD. The Survey has been IRB approved at Massachusetts General Hospital.

*Agreement of Subject I have read the above information and understand the terms of my participation. I agree to participate in this study.

- ◯ Yes
- ◯ No

2. What is your age (in years)

- 25-34
- 55-64
- 35-44
- 65+
- 45-54
- Other (please specify)

3. What is your level of training

- Resident
- Assistant Professor
- Fellow
- Associate professor
- Instructor
- Professor
- Other (please specify)

4. Gender identity

- Female
- Transmale
- Male
- Queer
- Transfemale
- Genderqueer
- Prefer to self describe

5. Are you Hispanic, Latino or Spanish origin?

- Yes
- No
- Other (please specify)

6. Are you:

- White
- Black or African American
- American Indian or Alaska Native
- Asian Indian
- Chinese
- Filipino
- Japanese
- Korean
- Pakistani
- Vietnamese
- Native Hawaiian
- Guamanian
- Chamorro
- Samoan
- Multi-racial
- Other (please specify)

7. What is your specialty?

8. Are you an international student/trainee or on a visa (i.e., currently on an F-1, J-1, H-1B, or another visa)

- Yes
- No
- Other (please specify)

9. Region of country

10. What was your year of graduation from your terminal training (ie clinical fellowship) before searching for a full-time position?

- prior to 2015
- 2015
- 2016
- 2017
- 2018
- 2019
- 2020
- 2021
- 2022
- 2023
- 2024
- N/A
- Other (please specify)

11. What is your PREFERRED research to clinical ratio (research % / clinical %)?

- 100/0
- 80/20
- 75/25
- 60/40
- 50/50
- 40/60
- 25/75
- 0/100
- N/A
- Other:

12. Why is this your PREFERRED research to clinical ratio?

13. What is your CURRENT research to clinical ratio (research % / clinical %)?

- 100/0
- 80/20
- 75/25
- 60/40
- 50/50
- 40/60
- 25/75
- 0/100
- N/A
- Other:

14. Does your current department/chair support your application to career development awards (like a NIH K award, DOD, foundation, specialty society)?

- Yes
- No
- Other:

15. Does your current department/chair provide equivalent base salaries between full time clinicians and physician-scientists?

- Yes
- No
- Unsure
- Other:

16. Does your current department/chair provide research incentives or research RVUs?

- ◯ Yes
- ◯ No
- ◯ Unsure

17. If yes to research RVUs, please provide examples of how this is awarded (Select all that apply)

- ☐ RVU credit for grant submissions
- ☐ Bonus compensation for funded grants
- ☐ Protected time tied to grant activity
- ☐ Departmental promotion points
- ☐ Other
- ☐ Annual productivity stipends
- ☐ Reduced clinical load or administrative responsibilities
- ☐ Recognition awards

18. Which NIH or VA career development award (CDA) have you applied for and/or received?

- ☐ K08 APPLIED
- ☐ K08 RECEIVED
- ☐ K99/R00 APPLIED
- ☐ K99/R00 RECEIVED
- ☐ K23 APPLIED
- ☐ K23 RECEIVED
- ☐ other K awards (K12, KL2, etc)
- ☐ Other (please specify)
- ☐ VA CDA: APPLIED
- ☐ VA CDA: RECEIVED
- ☐ DOD: APPLIED
- ☐ DOD: RECEIVED
- ☐ I have received a R01 or equivalent
- ☐ N/A

19. Were you funded by a foundation award/career development award (CDA) in your junior faculty position?

- ◯ Yes
- ◯ No
- ◯ NA

20. If you have a career development award, how many times did you have to apply for your current successful award?

- ◯ 1
- ◯ 2
- ◯ 3
- ◯ Other (please specify)
- ◯ ≥4
- ◯ NA

21. If you have a non NIH/VA CDA, what foundation or specialty society is your CDA from?

- ◯ Doris Duke
- ◯ Burroughs Wellcome Fund
- ◯ American heart association
- ◯ AACR
- ◯ ASCO
- ◯ IDSA
- ◯ ATS
- ◯ RSNA (Radiologic Society of North America)
- ◯ NA
- ◯ Other (please specify)

22. What is the length of the foundation/specialty society award in years?

- ◯ 1
- ◯ 2
- ◯ 3
- ◯ 4
- ◯ 5
- ◯ NA
- ◯ Other (please specify)

23. What is the award amount total?

24. Is the award mainly for salary?

- ◯ Yes
- ◯ No
- ◯ Other (please specify)

25. Is there protected research time restrictions for your foundation/specialty society award (>70% research requirement)?

- ◯ Yes
- ◯ No
- ◯ NA
- ◯ Other (please specify)

* 26. After you completed residency/fellowship, what was your first position?

- ◯ Academic center (research position)
- ◯ Academic center (clinical position)
- ◯ Academic center (hybrid research/clinical position)
- ◯ Industry (pharma, med tech, digital tech, etc)
- ◯ Other (please specify)
- ◯ Private practice
- ◯ An additional postgraduate residency or fellowship
- ◯ I'm still in my residency/fellowship

* 27. What are the top 3 challenges you have encountered thus far in your early faculty career/transition to early-career faculty?

- ☐ Lack of opportunity/funding
- ☐ Not finding a position in desired location
- ☐ Not able to find a desired position
- ☐ Loan repayment
- ☐ Salary disparity between my full-time clinical counterparts/Under-compensation
- ☐ Malpractice/lawsuit
- ☐ Discrimination/biases against your gender/ethnicity/sexual orientation
- ☐ Sexual harassment
- ☐ Balancing family and work responsibilities
- ☐ Balancing clinical, research, and educational responsibilities
- ☐ Satisfactory professional advancement
- ☐ Lack of support/resources by my chair/institution to apply for a career development award
- ☐ COVID-19 delaying/halting research projects
- ☐ Childcare
- ☐ Administrative burdens
- ☐ Hiring freezes
- ☐ Other (please specify)

* 28. What are the TOP TWO most important factors to you in selecting a position/career?

- [ ] Opportunities to do research
- [ ] Opportunities for patient care
- [ ] Opportunities for BOTH research and patient care
- [ ] Opportunities to teach
- [ ] Opportunities for community service
- [ ] Opportunities for interactions with trainees
- [ ] Ability to balance work and personal life
- [ ] Financial security
- [ ] Autonomy
- [ ] Prestige
- [ ] Childcare resources
- [ ] Childcare flexibilities
- [ ] Other (please specify)

* 29. In which area do you intend to spend the majority of your professional time? Please check the MOST likely/Highest priority one.

- ○ Education
- ○ Basic research
- ○ Clinical research
- ○ Translational research
- ○ Clinical duties
- ○ Therapeutics/diagnostics development
- ○ Advocacy
- ○ Administration
- ○ Other (please specify)

* 30. In the last six months, have you considered leaving academic medicine/research career within the next 2 years?

- ○ Yes
- ○ No
- ○ If considering leaving, please describe your top reasons for this:

* 31. In the next 5 years, how likely do you think it is that you will stay in academic medicine/research career?

- ○ 100%
- ○ >75%
- ○ 50-75%
- ○ 25-50%
- ○ <25%
- ○ Other (please specify):

* 32. Thinking about the reasons that might lead you to consider leaving your position, check all that apply

- ☐ I am unhappy, stressed, or otherwise less than satisfied with my current position.
- ☐ I am drawn to, excited by, or otherwise attracted to a different position.
- ☐ Burnout
- ☐ Funding challenges
- ☐ Undercompensation
- ☐ I would not consider leaving.
- ☐ Other (please specify):

* 33. Amid the recent policy changes affecting NIH funding and related research cuts, we have created the following section to assess how these decisions are affecting the research community, particularly early-career physician-scientists. Your responses will help inform efforts to advocate for sustained funding, institutional support, and career development resources.

Have you had an awarded NIH grant that was later rescinded?

- ○ Yes
- ○ No
- ○ NA
- ○ Other (please specify):

* 34. If yes, what type of grant was rescinded? (Select all that apply)

- [ ] R01
- [ ] R03
- [ ] R21
- [ ] R33
- [ ] R34
- [ ] F31
- [ ] F32
- [ ] K-series
- [ ] NA
- [ ] Other (please specify):

* 35. Was your NIH study section or advisory council meeting canceled or postponed?

- ( ) Cancelled
- ( ) Postponed
- ( ) No
- ( ) Not applicable
- ( ) Other (please specify):

* 36. What was the original date for your study section or advisory council meeting?

37. Has your study section or advisory council meeting been rescheduled?

- ( ) Yes
- ( ) No
- ( ) Not yet informed
- ( ) Not applicable

38. If your grant was paused, rescinded, or delayed, what were the primary consequences? (Check all that apply)

- [ ] Delay hiring
- [ ] Delay project
- [ ] Loss of momentum
- [ ] Reduced collaboration
- [ ] Loss of protected time
- [ ] No consequences
- [ ] NA
- [ ] Other (please specify)

39. Have the funding disruptions affected your mental or emotional well-being? (Select all that apply)

- [ ] Increased stress
- [ ] Burnout
- [ ] Depression symptoms
- [ ] Loss of motivation
- [ ] Uncertainty but hopeful
- [ ] No significant effects
- [ ] NA
- [ ] Other (please specify)

40. How would you describe your current outlook on your future as a physician-scientist?

- ○ Very hopeful
- ○ Somewhat hopeful
- ○ Neutral
- ○ Somewhat pessimistic
- ○ Very pessimistic
- ○ Other (please specify)

41. Do you plan to maintain, increase, or decrease research time in next 2–3 years?

- ◯ Maintain
- ◯ Increase
- ◯ Decrease
- ◯ Not sure
- ◯ Other (please specify)

[ ]

42. If planning to change research time, what are the primary reasons?

- ☐ · Funding loss
- ☐ · Institutional pressure
- ☐ · Unstable funding climate
- ☐ · Burnout
- ☐ · Lack of support
- ☐ · Job security
- ☐ Other (please specify)

[ ]

43. If considering reducing research time, what percentage reduction are you considering?

- ☐ <10%
- ☐ 10-25%
- ☐ 25-50%
- ☐ 50%
- ☐ >50%
- ☐ Other (please specify)

[ ]

44. What would help you maintain your current research effort in the face of funding or policy uncertainty? (Select all that apply)

- ☐ · Bridge funding
- ☐ · Transparent timelines
- ☐ · Institutional support
- ☐ · Grant mentorship
- ☐ · Alternative funding
- ☐ · Peer networks
- ☐ · Policy advocacy
- ☐ Other (please specify)

45. Are you concerned that future NIH funding constraints or policy shifts will disproportionately affect certain research areas?

- ○ Yes
- ○ No
- ○ Not sure
- ○ Other (please specify)

46. If yes, which areas are you most concerned about? (Select all that apply)

- ☐ · Vaccine research
- ☐ · LGBTQ health and rights
- ☐ · Diversity, equity, and inclusion (DEI) research
- ☐ · Reproductive health (including contraception and abortion)
- ☐ · Health disparity research
- ☐ · Climate and environmental research
- ☐ · Mental health
- ☐ Other (please specify)

47. What do you think are possible solutions? (check all that apply)

- ☐ · Public–private partnerships
- ☐ · Innovative financial models (e.g., sovereign wealth funds, private investment in biomedical research similar to Australia's Medical Research Future Fund)
- ☐ · Legislative advocacy and targeted policy changes
- ☐ · Institutional bridge funding or internal grant mechanisms
- ☐ · Multi-institutional research consortia for shared grant infrastructure
- ☐ · Increased role of philanthropy or foundation-based funding
- ☐ Other (please specify)

48. Have other countries reached out to try to recruit you for a research/clinical position?

- ◯ Yes
- ◯ No
- ◯ If yes, which country has tried to recruit you (please specify)

49. Have you considered leaving the US in order to continue your research career?

- ◯ Yes
- ◯ No
- ◯ If yes, which country (please specify)

50. Would joining an organization dedicated to both advocacy for funding, policy changes and provides career development content/networking for early-career physician-scientists be of interest to you?

- ◯ Yes
- ◯ No
- ◯ Not sure

51. Thank you for your time in helping to advocate for physician-scientists. Please let us know if you have any feedback, comments or suggestions
